# Training machine learning models on patient level data segregation is crucial in practical clinical applications

**DOI:** 10.1101/2020.04.23.20076406

**Authors:** Mustafa Umit Oner, Yi-Chih Cheng, Hwee Kuan Lee, Wing-Kin Sung

## Abstract

This article discusses the effect of segregation of histopathology images data into three sets; training set for training machine learning model, validation set for model selection and test set for testing model performance. We found that one must be cautious when segregating histological images data (slides) into training, validation and test sets because subtle mishandling of data can introduce data leakage and gives illusively good results on the test set. We performed this study on gene mutation prediction performance by using the deep neural network in the paper of Coudray et al. [1]. By using the provided code and the same set of data, we discovered that data segregation method of the paper suffered from a data leakage problem [2]. The paper pools all the slides from all patients and then segregates them exclusively into training, validation and test sets. In this way, none of the slides is used in more than one set. This seems to be a clean separation of the data. However, the paper did not consider that some slides were strongly correlated. For example, if the tumor of a patient is cut and stained to produce multiple slides, these slides are strongly correlated. If one slide is used for training and another one is used for testing, essentially, the deep neural network can memorize the pattern on the slide in the training set and apply this memory on the slide in the test set. Hence, by memorization, the deep neural network can predict very well on the slide in the test set. This mechanism of prediction is not useful in a practical clinical setting since no two tumors are the same in the real world. In this real setting, we demand the deep neural network to generalize across patients and tumors. Hereafter, we call this way of data segregation *slide-level segregation*. There is a better way to perform data segregation that is compatible for deployment of deep learning model in practical clinical settings. First, the patients are segregated exclusively into training, validation and test sets. All the slides belonging to the patients in the training set are used solely for training. Similarly, all the slides belonging to the patients in the test set are used for testing only. Segregation of data in this way forces the deep neural network to generalize across patients. We call this way of data segregation *patient-level segregation*.

In *slide-level segregation* approach analysis, we obtained similar results to that presented in the paper by Coudray et al. [1]: overall performance on the test set was good. However, it was illusory due to data leakage. The model gave very good testing results on the slides that come from a patient who also has slides in the training set. On the other hand, the test result was quite bad on the slides that come from a patient who does not have any slides in the training set. Hereafter, we call the slide in the test set as seen-patient data if the corresponding patient also has some slides in the training set. Otherwise, the slide in the test set is called unseen-patient data if the corresponding patient does not have slides in the training set. Furthermore, we analyzed performance of the model on the data segregated by the *patient-level segregation* approach. Note that, in this approach, all patients in the test set mimics the real world clinical workflow. We observed a significant drop in the performance of the model on the test set of *patient-level segregation* approach compared to the performance on the test set of *slide-level segregation* approach. Moreover, the performance of the model on the test set of *patient-level segregation* approach was very similar to the performance on the unseen-patients data in the test set of *slide-level segregation* approach. Hence, we conclude that *patient-level segregation* approach is crucial and appropriate to simulate real world scenario, where each patient in the test set can be thought as a patient walking into clinic tomorrow.

## Analysis Setup

In the paper “Classification and mutation prediction from non–small cell lung cancer histopathology images using deep learning” by Coudray et al. [1], it is claimed that six gene mutations in lung adenocarcinoma patients from TCGA Research Network (https://www.cancer.gov/tcga) can be predicted from histopathology slides. We downloaded the code from the github page link given in the paper, and downloaded the same slides from the TCGA data portal. Note that, in TCGA cohort, multiple slides are obtained from neighboring regions of the same tumor sample of a patient [3] (see Supp. Figure 1). Then, we conducted two different analysis by segregating the available data into training, validation and test sets by (i) *slide-level segregation* and (ii) *patient-level segregation* approaches (see Supp. Figure 2).

**Figure 1:**
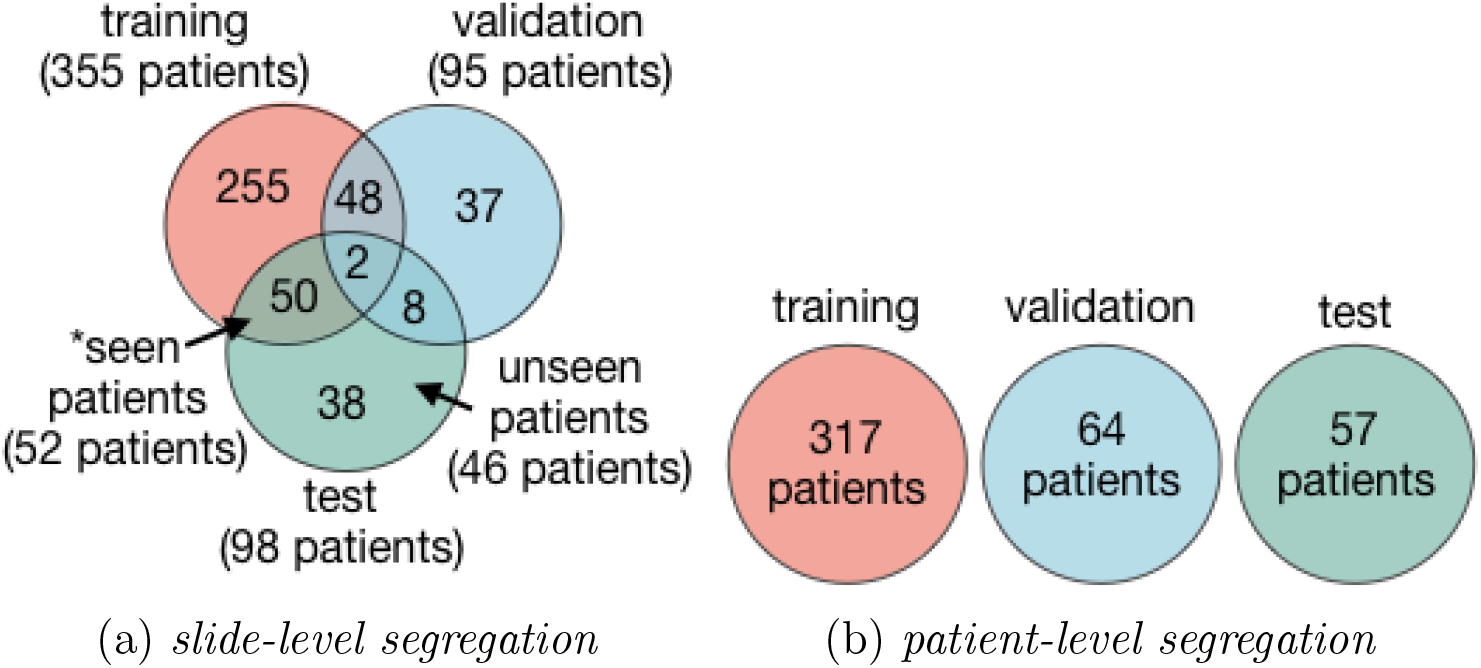
Number of patients in training, validation and test sets for (a) *slide-level segregation* and (b) *patient-level segregation* approaches. There are 438 patients in total. While all three sets are mutually exclusive in *patient-level segregation* approach, there are overlaps among sets in *slide-level segregation* approach. For example, as shown in the set marked by *, slides of 52 patients are used in both training and test sets.

**Figure 2:**
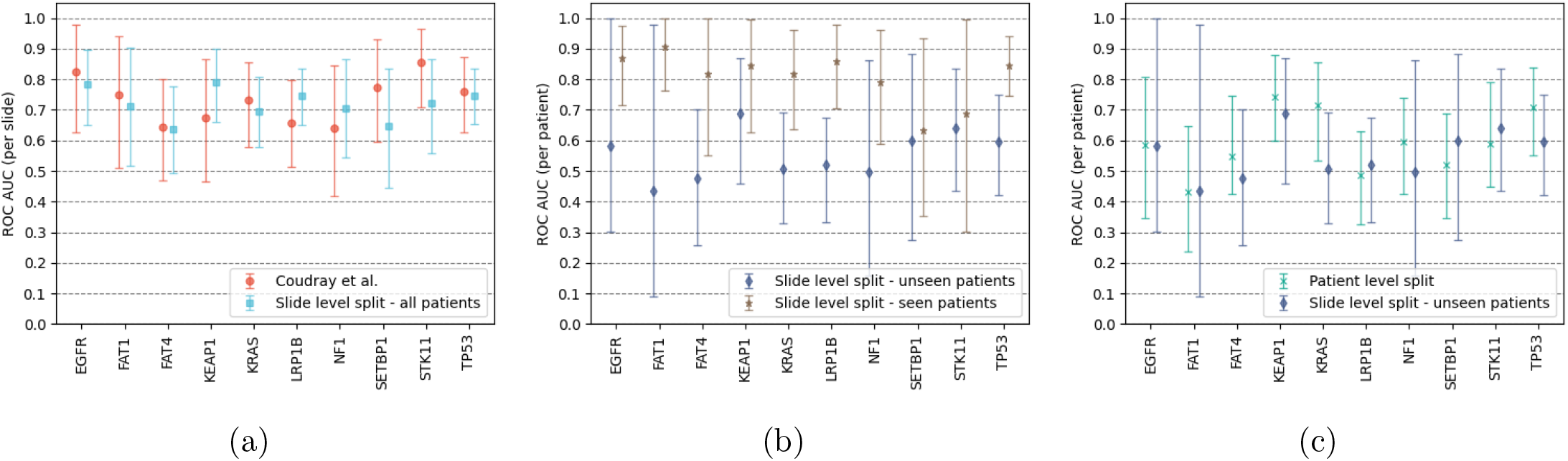
Comparison of area under receiver operating characteristics curve (ROC AUC) values with 95% confidence intervals on test sets. While the markers show mean values, upper and lower bars on the plots show upper and lower bounds of the 95% confidence intervals. (a) The results given in the paper of Coudray et al. [1] and the results obtained in our analysis with *slide-level segregation* approach are presented. All patients in the test set were used during analysis. (b) Comparative ROC AUC results for unseen-patients and seen-patients data of the test set of *slide-level segregation* approach are shown. (c) Comparative ROC AUC results for the test set of *patient-level segregation* approach and unseen-patients data of the test set of *slide-level segregation* approach are shown. Note that ROC AUC (per slide) and ROC AUC (per patient) in the y-axis indicate that ROC curves were obtained over slide level predictions and patient level predictions, respectively.

### Reproducing the results of the paper: *slide-level segregation*

In Coudray et al. [1], *slide-level segregation* approach was used in the data segregation. By using the same approach, slides of the 438 patients were assigned to training, validation and test sets (see Supp. File 1). The number of patients in each set is shown in Figure 1a. Overlapping regions among the sets show number of patients, whose data shared by multiple sets. For example, slides from 52 patients were assigned to both training and test sets. The slides of these 52 patients in the test set were the seen-patients data. Similarly, the slides of the other 46 patients in the test set were the unseen-patients data.

Area under receiver operating characteristics curve (ROC AUC) values with 95% confidence intervals were obtained at the end of the experiment. The results obtained from the experiment and that from the paper [1] are presented in Figure 2a. For most of the genes, the results from our experiment are very close to the results in Coudray et al. The small deviation was due to randomness involved in initialization of training of the machine learning model and handling of data. Hence the results in Coudray et al. were reproduced successfully.

Due to the data leakage issue, performance of the model may be higher on seen-patients data. In order to check if this is true, a comparative analysis on unseen-patients data and seen-patients data was conducted. Results of the analysis are presented in Figure 2b. As a prominent result of the *data leakage problem*, there is a significant performance difference between the unseen-patients data and seen-patients data for almost all of the genes. The performance on all patients data resides in between the performances on unseen-patients data and seen-patients data.

### Simulating the real world scenario: patient-level segregation

In order to simulate real world scenario, *patient-level segregation* approach must be employed so that each patient in the test set can be thought as a patient walking into clinic tomorrow. Slides of the 438 patients were assigned to training, validation and test sets by using this approach (see Supp. File 2). Number of patients in each set is shown in Figure 1b.

The results of the analysis are summarized in Figure 2c together with the results obtained on unseen-patients data of the test set of *slide-level segregation* approach. As anticipated, the results are quite similar to each other for almost all of the genes since none of the slides of the patients in these sets had been seen by the deep neural network during training.

Lastly, the performance obtained on the test set of *patient-level segregation* approach (see Figure 2c) is significantly different than the performance obtained on the test set of *slide-level segregation* approach (see Figure 2b).

## Conclusion

We showed that *slide-level segregation* approach has data leakage problem and does not mimic the real world clinical workflow. On the other hand, *patient-level segregation* approach is both appropriate and crucial to segregate data for training machine learning models to be used in practical clinical applications.

## Data Availability

All data is available online at TCGA Data Portal.

https://www.cancer.gov/tcga

## Acknowledgements

This work is partly supported by the Biomedical Research Council of the Agency for Science, Technology, and Research, Singapore and the National University of Singapore, Singapore.

## Author Contributions

M.U.O., H.K.L. and W.K.S. designed the experiments; Y.C.C. run the experiments; M.U.O. analyzed the results and drafted the manuscript; H.K.L. and W.K.S. substantively revised the manuscript.

## Supplementary Materials

**Supplementary Figure 1.**
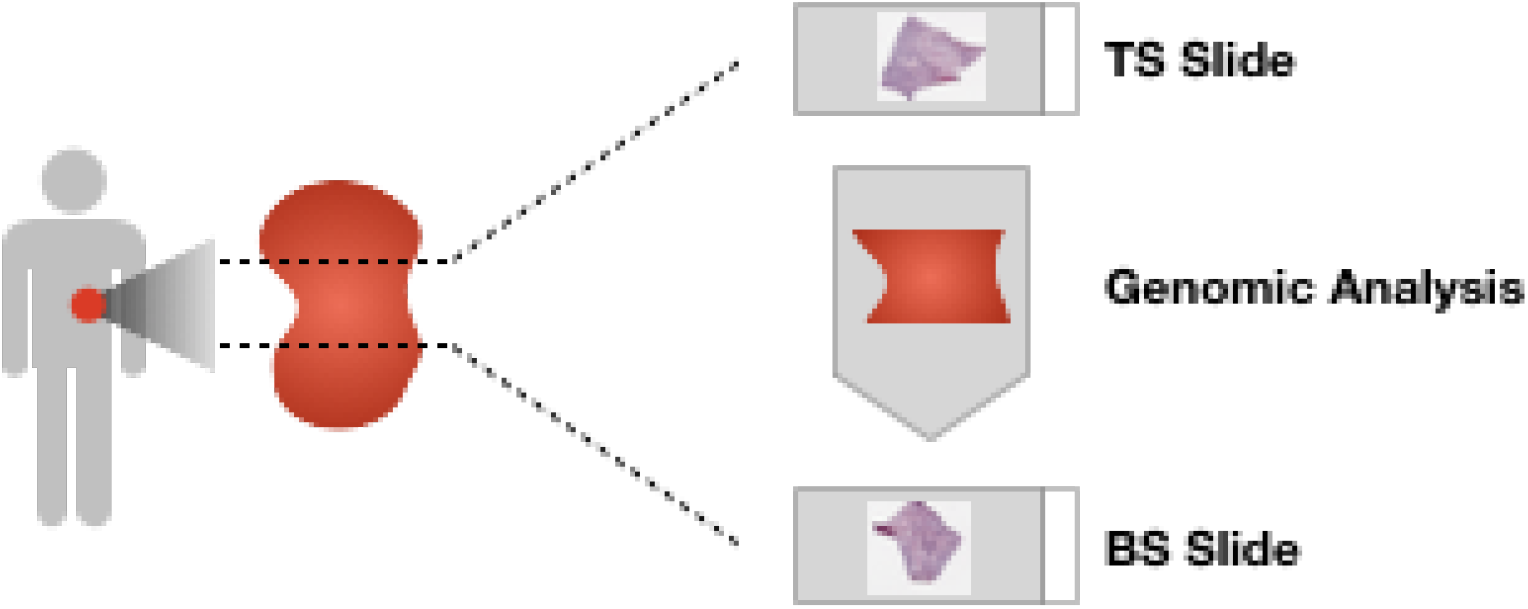
In TCGA cohort, multiple histopathology slides are obtained from a sample of a patient. A sample is obtained from surgical resection. While the ‘top-section’ (TS) and ‘bottom-section’ (BS) are used to prepare ‘TS’ and ‘BS’ slides, the middle portion of the sample is used for genomic analysis. Note that this figure is adapted from [3].

**Supplementary Figure 2.**
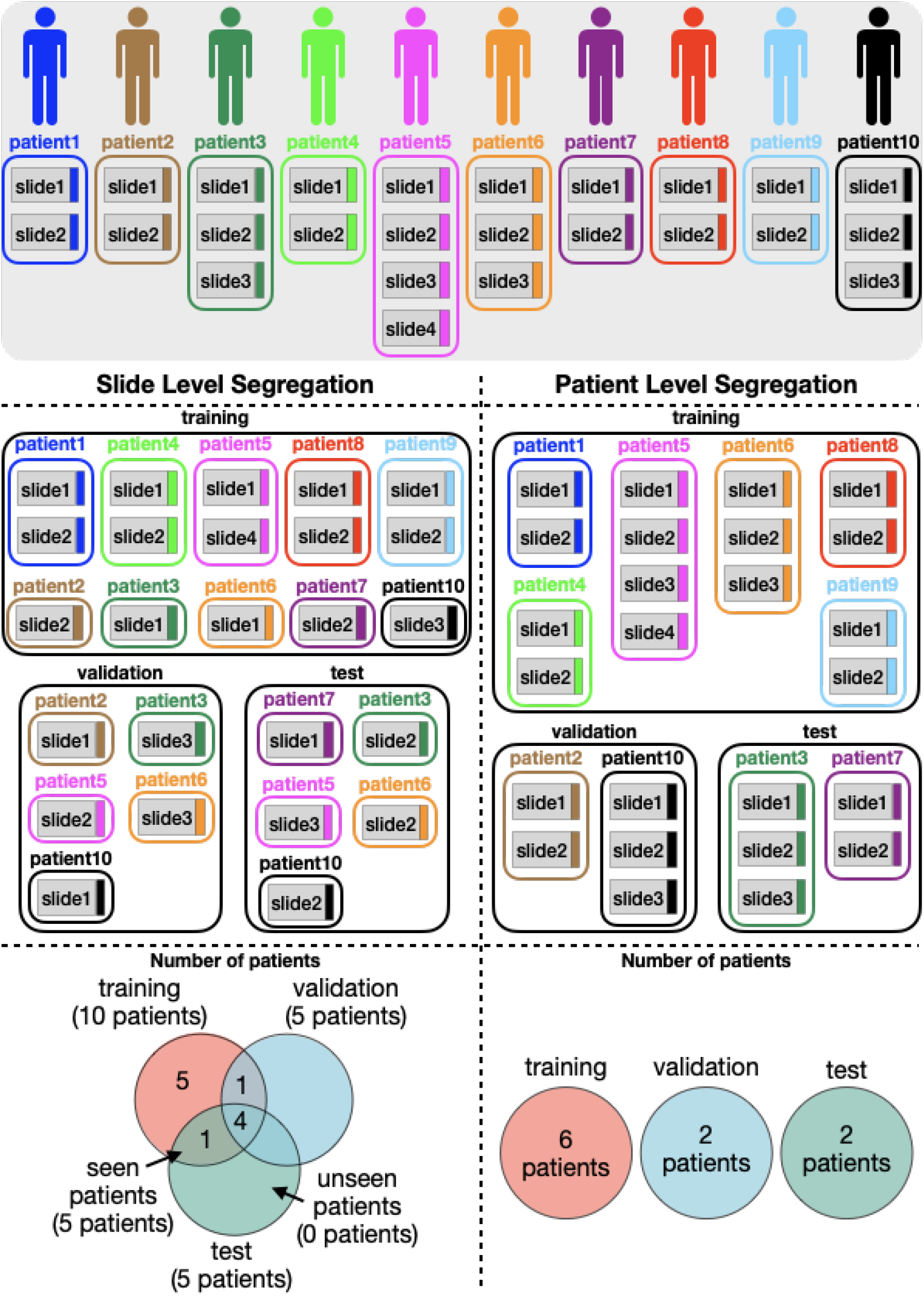
Below figure illustrates *slide-level segregation* approach and *patient-level segregation* approach on an example dataset with 10 patients. In *slide-level segregation* approach, each slide is assigned to either one of the training, validation or test sets. Although the slides in training, validation and test sets are different, the slides that come from the same patient in these sets are highly correlated. Hence, this results in a serious data leakage problem. For example, while ‘slide1’ and ‘slide4’ of patient5 are assigned to the training set, ‘slide2’ is assigned to the validation set and ‘slide3’ is assigned to the test set. On the other hand, in *patient-level segregation* approach, all the slides of a patient are assigned to either one of the training, validation and test sets. Therefore, all the sets are from independently sampled patients and there is no data leakage. Number of patients in training, validation and test sets are also shown at the bottom of the figure for both of the approaches. While there is no seen-patient in the test set of *patient-level segregation* approach, there are seen-patients in the test set of *slide-level segregation* approach. For this example, 5 seen-patients were in the test set of *slide-level segregation* approach.

### Supplementary File 1

“slide_level_segregation_approach.xlsx”

The list of patient ids and slide ids in training, validation and test sets obtained by *slide-level segregation* approach are given. Seen-patients in the test set are highlighted inside the file.

### Supplementary File 2

“patient_level_segregation_approach.xlsx”

The list of patient ids and slide ids in training, validation and test sets obtained by *patient-level segregation* approach are given.

